# A rare missense variant in the *ATP2C2* gene is associated with language impairment and related measures

**DOI:** 10.1101/2021.01.12.21249315

**Authors:** Angela Martinelli, Mabel Rice, Joel B. Talcott, Rebeca Diaz, Shelley Smith, Muhammad Hashim Raza, Margaret J. Snowling, Charles Hulme, John Stein, Marianna E. Hayiou-Thomas, Ziarih Hawi, Lindsey Kent, Samantha J. Pitt, Dianne F. Newbury, Silvia Paracchini

## Abstract

At least 5% of children present unexpected difficulties in expressing and understanding spoken language. This condition is highly heritable and often co-occurs with other neurodevelopmental disorders such as dyslexia and ADHD. Through an exome sequencing analysis, we identified a rare missense variant (chr16:84405221, GRCh38.p12) in the *ATP2C2* gene. *ATP2C2* was implicated in language disorders by linkage and association studies, and exactly the same variant was reported previously in a different exome sequencing study for language impairment (LI). We followed up this finding by genotyping the mutation in cohorts selected for LI and comorbid disorders. We found that the variant had a higher frequency in LI cases (1.8%, N=360) compared to cohorts selected for dyslexia (0.8%, N = 520) and ADHD (0.7%, N = 150), which presented frequencies comparable to reference databases (0.9%, N = 24,046 gnomAD controls). Additionally, we observed that carriers of the rare variant identified from a general population cohort (N=42, ALSPAC cohort) presented, as a group, lower scores on a range of reading and language-related measures compared to controls (N=1825) (minimum p = 0.002 for nonword reading). *ATP2C2* encodes for an ATPase (SPCA2) that transports calcium and manganese ions into the Golgi lumen. Our functional characterization suggested that the rare variant influences the ATPase activity of SPCA2. Thus, our results further support the role of *ATP2C2* locus in language-related phenotypes and pinpoint the possible effects of a specific rare variant at molecular level.

## INTRODUCTION

Speech and language disorders represent one of the most common childhood disabilities and account for a large proportion (23%) of paediatric referrals and statements of special educational need (School census, England, January 2019, https://www.gov.uk/government/statistics/special-educational-needs-in-england-january-2019).

Speech and language deficits are often the secondary manifestations of a pre-existent neurological or medical condition (e.g. Down syndrome, intellectual disability, autism spectrum disorder, hearing loss, traumatic brain injury and mental retardation). Severe and persistent language difficulties can be observed also despite normal intelligence and access to adequate educational opportunities. In this case children were diagnosed with specific language impairment (SLI) (1). Recently, the use of this term has been replaced by the developmental language disorder (DLD) definition (2,3) which captures a wider group of children presenting co-occurring conditions such as intellectual disability, ADHD and developmental coordination disorder (DCD). Previous genetic research, and the samples described here, refer to participants recruited under the SLI criteria and therefore we will use the terms SLI and LI hereafter.

SLI affects up to 8% of pre-school children (4,5) and has a significant impact on social, academic and psychological outcomes (6–10). SLI often co-occurs with other neurodevelopmental disorders such as dyslexia (11,12) and attention deficit/hyperactivity disorder (ADHD) (13). The comorbidity observed between these conditions suggests that SLI, dyslexia and ADHD may be different manifestations of the same cognitive deficit or share common genetic factors (14). This hypothesis is supported by significant genetic overlap across studies as demonstrated by a recent genome-wide association study (GWAS) for dyslexia which showed significant shared contributions across disorders (15).

A strong genetic component, of ∼ 70%, was first proposed by twin and familial studies (16–20). Initially, linkage studies conducted across multiplex families led to the identification of 5 susceptibility loci on chromosomes 2q36 (SLI5, OMIM #615432) (21), 7q35-q36 (SLI4—OMIM #612514) (22), 13q21 (SLI3—OMIM #607134) (23,24), 16q (SLI1—OMIM #606711) and 19q (SLI2— OMIM #606712) (25–28). Fine mapping within these regions reported associations of common variants at the *ATP2C2, CMIP, CNTNAP2* genes with quantitative measures of language abilities (22,29), including the phonological memory nonword repetition task (NWR), which is considered as a strong endophenotype for SLI (30).

GWAS for language skills have been conducted in clinical and epidemiological samples, but have been limited to a few studies of relatively small sample sizes (31). In the first case, the only locus reaching genome-wide significance in SLI individuals was found in the *NOP9* gene, when including parent-of-origin effects (32,33). In the second case, the most significant result was the association between the locus 3p12.3, near *ROBO2*, and expressive vocabulary in the early phase of language acquisition (34). A GWAS conducted in two population cohorts reported an association close to statistical significance between *ABCC13* gene and the NWR test (35). Other studies combined analyses for language and reading abilities using both a case-control (36) and quantitative design (37) reporting only marginally significant associations.

Chromosomal rearrangements and copy number variants (CNV) have also been implicated in the aetiology of language impairment. Children with SLI show a higher frequency of sex chromosome aneuploidies (38), and an increased CNV burden compared to population controls (39,40). A balanced translocation interrupting the gene *SEMA6D* (41) has been identified in a child with SLI. Rare deletions have been reported in the *ZNF277* (42) and *TM4SF20* (SLI5) genes (21). Larger *de novo* deletions have been reported across the *ATP2C2* gene in a 10 year old boy with receptive and expressive language delay (43) and at chromosome 15q13.1–13.3 in a child with LI (44). Single locus mutations have been implicated in severe forms of language disorders (31,45). De novo mutations in the *FOXP2* gene cause childhood apraxia of speech (CAS), a severe oral-motor delay accompanied by deficits in expressive and receptive language (46).

Next-generation sequencing (NGS) technology is uncovering the role of rare variants in complex traits. NGS in individuals with CAS or SLI pinpointed to novel pathogenic variants in coding and non-coding regions (45,47,48). Whole exome sequencing (WES) analysis in 43 unrelated probands affected by severe SLI (49) reported several risk variants but suggested that a single mutation might not be sufficient to lead to a disorder unless in combination with other factors in line with previous studies (50–52).

Here, we followed up a specific finding from ongoing exome sequencing analyses on large pedigrees selected for having multiple members affected by SLI. We identified, in one family, a rare non-synonymous variant in the *ATP2C2* gene (Chr16:84405221 GRCh38.p12, rs78887288, NM_001286527:exon3:c.G304A:p.V102M), previously implicated in SLI through association studies (29,43)(29,43). Exactly the same variant, was also reported in the WES study by Chen and collaborators (49) bolstering the idea that this rare variant has a role in susceptibility to language disorders. Nonetheless, given its rarity, larger screens and functional studies are required to support this role.

We screened the chr16:84405221 variant in larger cohorts of children with language difficulties. Due to the high rate of comorbidity between SLI, dyslexia and ADHD, the screening included cohorts selected for these conditions.

We found that this variant had a higher frequency in the SLI and LI cases compared to the control group and cohorts selected for dyslexia or ADHD. Additionally, this variant was associated with low performance in a range of reading and language measures. By means of a colorimetric ATPase assay, we further investigated the effects of the variant on the function of SPCA2, the calcium and manganese transporter encoded by *ATP2C2*. The mutation displayed a lower, albeit not significant, ATPase activity compared to SPCA2 WT.

## MATERIAL AND METHODS

### Discovery pedigree

An exome sequencing analysis was conducted in six pedigrees of SLI probands with multiple affected members per family, selected from an ongoing longitudinal study in the Midwestern United States (53,54). The full results of this screening will be reported in a separate manuscript. SLI probands were identified based on language performance 1 standard deviation (SD) or more below the mean on an age appropriate omnibus language test (53), and met the following entrance screening criteria: (i) a standard score above 85 on an initial age appropriate test of non-verbal IQ, (ii) normal hearing acuity, (iii) no history of neurological disorders or diagnosis of autism, and (iv) intelligible speech sufficient for language transcription. Except for normal hearing acuity, the entrance screening criteria did not apply to siblings and parents of SLI probands, as their intellectual status was an outcome of the study (53).

Here we report the analysis and follow up of a specific variant of interest (Chr16: 84405221) in one family (Figure 1). A male SLI proband, his younger male sibling (Sib 1) and their parents were selected for WES. All family members including the other two siblings were genotyped for the *ATP2C2* variant. Longitudinal phenotype data on the proband were assessed from age 6 to 19 years and the siblings were assessed also at multiple time points. Both mother and father were tested at a single time point. All siblings and the father met the criteria above for a clinical diagnosis of SLI. The sibling selected for WES was also delayed on the Test of Early Grammatical Impairment (TEGI) until age 7 years, 6 months (55). The children were also delayed in receptive vocabulary and oral reading across age, as well as in mean length of utterance when young. The study was approved by the institutional review boards at the University of Kansas and at the University of Nebraska Medical Center. Appropriate informed consent was obtained from the subjects and/or their caregivers.

**Figure 1.**
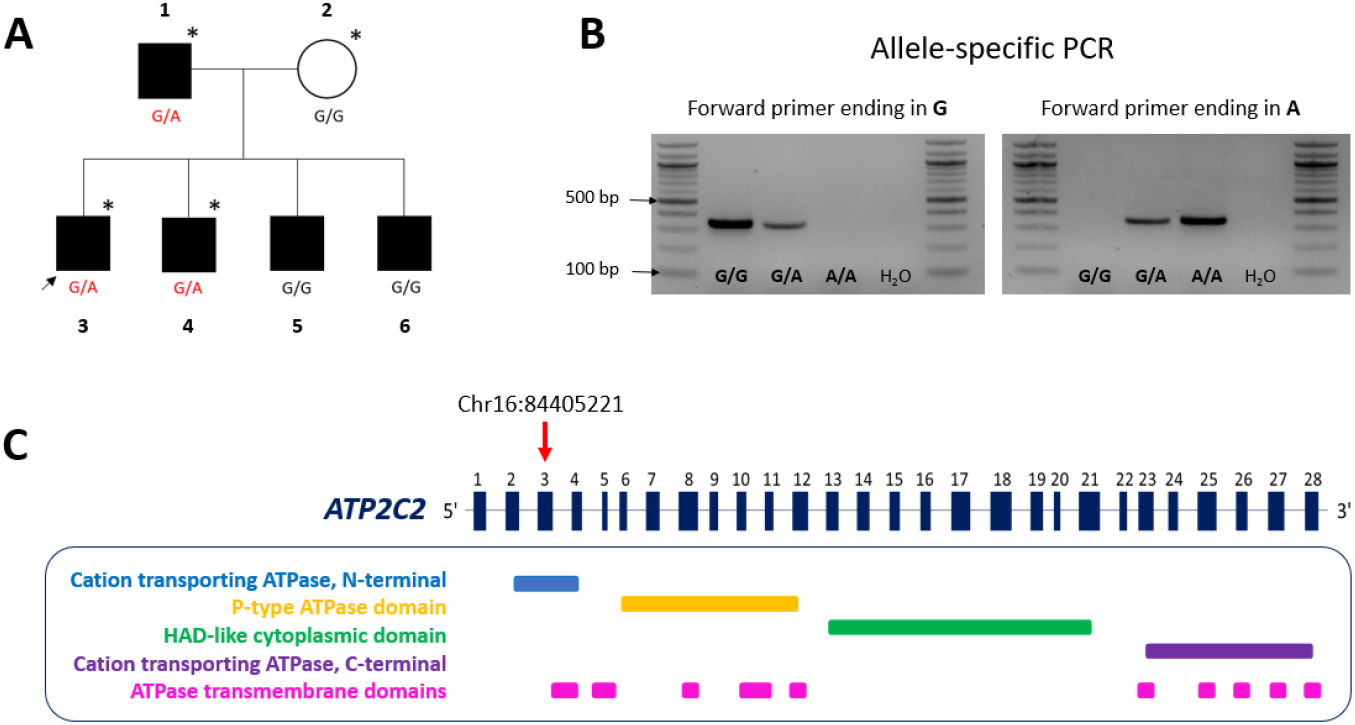
The rare *ATP2C2* missense variant (Chr16:84405221). **A**) Discovery pedigree. Father (1), mother (2), proband (3) and siblings (4, 5, 6). Subjects with SLI are shaded black and unaffected are white. Individuals included in the WES screening are indicated by an asterisk. **B**) Allele-specific PCR. The presence of the missense variant was validated in follow up screening using PCR 3’-end mismatch primers. Depending on the allele present, the forward primer anneals perfectly to the template or forms a mismatch. The reaction gives an amplicon only in the first case. **C**) On top, a schematic of the *ATP2C2* gene. Exons are represented by numbered boxes. The rare variant is localised in the third exon (red arrow) and causes the substitution of the residue from valine to methionine. Exons and introns of *ATP2C2* gene are not to scale. At the bottom, predicted domains of the protein (http://www.rcsb.org/pdb/protein/O75185) (82). The variant changes the amino acid sequence which codes for the cation transporting ATPase domain at the N-terminal of the protein.

### Validation, genotyping and bioinformatics prediction for the variant

Sanger sequencing was used to validate the variant and test all available family members in the discovery pedigree.

The variant was then screened in larger cohorts with a TaqMan™ SNP Genotyping Assay run on a ViiA7 instrument (Life Technologies, Paisley, UK). The presence of the mutation was validated using PCR 3’-end mismatch primers (Figure 1B). All primer sequences are reported in Supplementary Table S1. For protocol details see Supplementary Table S2 and S3, and Supplementary Figure S1. The functional impact of the rare *ATP2C2* variant was estimated using the default settings of seven different *in silico* prediction algorithms (Table 1), SIFT (56), Mutation Assessor (57), PROVEAN (58), PolyPhen-2 (59), MutPred (60), INPS-MD (61), MutationTaster (62).

**Table 1.** Prediction of the possible effects of the *ATP2C2* missense variant (chr16:84405221).

| Computational tool |  |  |  | ATP2C2 variant |  |
| --- | --- | --- | --- | --- | --- |
| Name | Effect | Score cut-off | Result | Score | Prediction |
| SIFT | activity | $0 < s < 0.05$ | Deleterious | 0.24 | tolerated |
| Mutation Assessor | activity | $s > 3.5$ | high functional impact | 1.535 | low functional impact |
| PROVEAN | activity | $s \leq -2.5$ | Deleterious | -0.399 | neutral |
| PolyPhen-2 | pathogenicity | $0.85 < s < 1.0$ | Damaging | 0.002 | benign |
| MutPred | pathogenicity | $s > 0.50$ | Pathogenic | 0.114 | benign |
| INPS-MD | stability<br>(Gibbs free energy) | $s < -0.5$ | large destabilisation | -0.073 | weak destabilising effect |
| MutationTaster | structure | $s \geq 151$ | radical substitution | 21 | Conservative |

### Cohorts for follow-up analysis

The genotyping was conducted across different UK cohorts of children with language impairment, dyslexia or ADHD used in previous genetic studies (Table 2). We use the SLI term when participants were recruited under SLI criteria and the term LI to refer to individuals who did not receive a specific diagnosis but presented language difficulties on psychometric tests. Assignment to a case group was based either on existing clinical diagnoses (York and ADHD cohorts) or derived from poor performance on different psychometric tests as specified below. Exclusion criteria included non-European ethnicity, English as a second language, signs of other sensory or neurological conditions and missing information for the selected phenotypes.

**Table 2.** Frequency of the rare allele A (Chr16:84405221) across different cohorts.

| Cohort | Cohort type | N | Allele A count | Allele A frequency | Reference |
| --- | --- | --- | --- | --- | --- |
| SLIC | Clinical | 161 cases <sup>(1)</sup> | 7 | 2.2% | (26) |
| York | Longitudinal | 23 LI cases<sup>(2)</sup> | 0 | 0% | (64) |
|  |  | 18 dyslexia cases | 3* | 8.3% |  |
|  |  | 72 TD | 1 | 0.7% |  |
| Dyslexia | Clinical | 471 cases | 5 | 0.5% | (71) |
| ADHD | Clinical | 141 cases | 2 | 0.7% | (72) |
| ALSPAC | Longitudinal Epidemiological | 170 LI cases<sup>(3)</sup> | 5 | 1.5% | (74,75) |
|  |  | 31 dyslexia cases | 0 | 0% |  |
|  |  | 10 ADHD cases | 0 | 0% |  |
| Combined cases |  |  |  |  |  |
| Combined SLI/LI samples |  | 360 cases<sup>(4)</sup> | 13 | 1.8% |  |
| Combined dyslexia samples |  | 520 cases | 8* | 0.8% |  |
| Combined ADHD samples |  | 151 cases | 2 | 0.7% |  |
| Controls |  |  |  |  |  |
| ALSPAC | Longitudinal Epidemiological | 681 controls | 14 | 1% | (74,75) |
| European population (gnomAD v2.1.1) |  | 24046 controls | 426** | 0.9% | (79) |
| gnomAD v2.1.1 |  | 59718 controls | 584*** | 0.5% |  |

\* includes N=1 homozygote; \*\* includes N=2 homozygotes; \*\*\* includes N=3 homozygotes
**(1)** including the probands analysed by Chen and colleagues through exome sequencing (Chen et al., 2017);
**(2)** includes N=11 individuals with comorbid SLI and dyslexia; **(3)** includes N=9 individuals with comorbid LI and dyslexia
**(4)** including the 6 probands screened by whole exome sequencing
**Abbreviations:** ADHD: attention deficit hyperactivity disorder; ALSPAC: Avon Longitudinal Study of Parents and Children; SLIC, specific language impairment Consortium; TD: typically developing

The SLI Consortium (SLIC) cohort consisted of 263 families (263 probands, 258 siblings, 551 parents) (age range 5-19 years) recruited across different UK clinics (29). Participants were assigned to a case category when they scored ≥ 80 on performance IQ (PIQ) and had impaired expressive or receptive language skills (ELS or RLS score ≥ 1.5 SD below the normative mean of the general population) as assessed through the Clinical Evaluation of Language Fundamentals test (CELF-R, revised version) (63). If the same family comprised several cases, the most severe sibling was included into the analysis. Severity was determined by comparing the scores for ELS and RLS composite measure. If the cases performed at the same level on these measures, inclusion was determined by the most severe score on the NWR task. Overall, 137 unrelated cases met these criteria. A subset of this sample was used for the WES analysis conducted by Chen and colleagues where the rare variant was also reported (49). The ethical approval was given by local ethics committees of the hospitals involved in the consortium, and all subjects provided informed consent.

The York cohort consisted of 115 families (115 probands, 27 siblings, 184 parents), collected for a longitudinal study designed to investigate language and reading development (64). Participants entered the study for either having a family history of dyslexia or showing preschool language difficulties. A third group was recruited as control or typically developing children (TD). At 8 years the children were given a clinical diagnosis for SLI (N = 23, including 11 individuals with dyslexia) and dyslexia (N = 18). Ethical approval was obtained from the Yorkshire & The Humber - Humber Bridge NHS Research Ethics and the Ethics Committee at the University of York - Department of Psychology. Informed consent was given by all the participants and/or their parents.

Participants were assigned to a category of dyslexia from two other separate cohorts of families and individual cases. The criteria for inclusion were PIQ ≥ 85 and a score of 1 SD below the population mean on the single-word reading (READ) test measured through the British Ability Scales (BAS) (65) or the Wide Range Achievement Test (WRAT-R, revised version) (66). The first cohort comprised 290 families (689 siblings, 580 parents) (age range 6–27 years) described before (67). All the subjects were recruited by the Dyslexia Clinic at the Royal Berkshire Hospital, Reading, United Kingdom. When multiple children from the same family met the above criteria, we referred to other psychometric measures and selected the individual with the most severe phenotype on the following tasks: BAS single-word spelling accuracy (SPELL) (65), phonological decoding ability (CCN) (68), phonemic awareness (SPOON) (69) and orthographic coding skill assessed through the irregular word (CCI) (68) or forced word choice test (OLSON) (70). The Dyslexia Cases cohort, N = 592 singletons (age range 6–18 years), was collected from either the Dyslexia Research Centre clinics in Oxford and Reading, or the Aston Dyslexia and Development Clinic in Birmingham (71). In total, N = 471 participants met the above criteria for dyslexia. The study was approved by the Oxfordshire Psychiatric Research Ethics Committee and the Aston University Ethics Committee and informed consent was given by all the participants and/or their caregivers.

The ADHD cohort included 159 trios (age range 5-16 years) recruited from child psychiatry clinics in United Kingdom and Ireland (72). Participants were interviewed employing the Child and Adolescent Psychiatric Assessment (CAPA) (73). All the probands fulfilled the DSM-IV (American Psychiatric Society, 1994) diagnostic criteria for ADHD. Ethical approvals were acquired from the Trinity College Dublin in Ireland, and the University of Birmingham in UK. Informed consent was obtained from the parents of all the participants.

In addition to the clinical cohorts, we analysed a subset of samples from the Avon Longitudinal Study of Parents and Children (ALSPAC, see Supplementary material) (74,75) for which whole-genome (WGS) sequence data were generated as part of the UK10K project (76). Participants were excluded if they reported a non-white European ethnicity, presented with overt hearing difficulties or were affected by neurological conditions which may explain their language difficulties, such as autism, Asperger syndrome or pervasive developmental disorder. Individuals with missing information for the phenotypes of interest were also excluded.

The criteria for an assignment to a LI category were a PIQ ≥ 80 at age of 8.5 and a score either 1 SD below the population mean on the Wechsler Objective Language Dimensions (WOLD) comprehension test (77) at 8 years, or 1 SD below the population mean on both tests of syntax, and intelligibility and verbal fluency, as assessed through the Children’s Communication Checklist (CCC) (78) at 7.5 years. Assignment to the dyslexia category was given based on a diagnosis reported by the mother through a questionnaire and a PIQ score higher or equal to 85 at the age of 8.5. Assignment to the ADHD category was given if the subject met the DAWBA DSM-IV diagnostic criteria and had a PIQ > 80 at the age of 8.5. Please note that the ALSPAC study website contains details of all the data that is available through a fully searchable data dictionary and variable search tool (https://www.bristol.ac.uk/alspac/researchers/our-data/).

Participants were assigned to a ‘super-control’ group (N=681) when they performed above the mean on tests of receptive (WOLD comprehension) and expressive (CCC syntax and intelligibility and fluency scores) language and had both verbal and non-verbal IQ ≥ 80. From the controls we also excluded any child who self-reported a need for special educational support or speech and language therapy and children who self-reported a diagnosis of neurodevelopmental disorder (ADHD, conduct disorder, anxiety disorder, depressive disorder, dyslexia, dyspraxia, dyscalculia, learning difficulties, speech and language difficulties or developmental delay).

Overall, in the ALSPAC sample, we identified: N = 170 LI cases, N = 31 dyslexia cases, N = 10 ADHD cases and N = 681 controls. Individuals with comorbid LI and dyslexia were included in the LI category.

Ethical approval for the study was obtained from the ALSPAC Law and Ethics Committee and the Local Research Ethics Committees. Consent for biological samples has been collected in accordance with the Human Tissue Act (2004). Informed consent was signed by the parents after having received a complete description of the study at the time of enrolment into the ALSPAC project, with the option for them or their children to withdraw at any time.

The gnomAD v2.1.1 database (https://gnomad.broadinstitute.org/) was used as a reference European population (79). Of note, this source includes the UK10K samples.

### Quantitative analysis

While in most cohorts the number of carriers of the rare variant was too small to conduct any meaningful quantitative analysis, the ALSPAC dataset was suitable for this option. All the individuals with available genotype (N = 1867) were included into the analysis. Carriers (N = 42) and non-carriers (N = 1825) were compared for the mean score calculated for reading and language-related measures including: single-word reading accuracy (READ, target age: 7.5 year), single-word spelling accuracy (SPELL, 7.5 year), phoneme awareness (PHONEME, 7.5 year), working memory (MEMSPAN, 10.5 year), listening and comprehension test (WOLD, 8.5 year), phonological short-term memory test (NWR, 8.5 year), single-non-word reading accuracy (NW-READ, 9.5 year), average of the first seven scales from the Children’s Communication Checklist (CCC-average7, 7.5 year) and performance IQ (PIQ, 8.5 year) (See Supplementary Table S4 for more details). Statistical significance was calculated with the Student’s t-test (equal variances, two-tailed distribution).

### ATPase assay

The human *ATP2C2* coding sequence was cloned into the expression vector pcDNA™5/TO (Invitrogen, kind gift from Professor Galione (80). *ATP2C2* cDNA was obtained from SK-BR-3 cells maintained under standard conditions and amplified by high fidelity PCR. The primers were designed to include a *HindIII* site prior the Kozak consensus sequence and a *NotI* site immediately before the stop codon (See Supplementary Table S1 for the primer sequences). The *ATP2C2* open reading frame was placed under the control of a constitutive CMV promoter and in frame with a downstream HA epitope tag. The SPCA2-HA tagged plasmid was modified to introduce the missense variant (allele A). The mutated construct (SPCA2 p.V102M) was built using the GeneArt™ Site-Directed Mutagenesis kit (Thermo Fisher) according to the manufacturer’s instructions. The sequences were verified by Sanger sequencing (See Supplementary Table S1 for primer sequences).

Microsomes were prepared as described by Vandecaetsbeek (81) with minor modifications (See Supplementary material for details). HEK293 cells were transfected with the *ATP2C2* expressing constructs. The empty vector and untransfected cells were used as controls.

ATPase activity was evaluated using the P_i_ColorLock™ Gold kit (Expedeon, Abcam) according to the manufacturer’s specifications. The assay was conducted in a 96-well plate and each sample run in duplicates. The amount of free inorganic phosphate (Pi) released from each reaction was calculated by plotting the absorbance against a series of Pi standards of known concentration.

The data generated were presented as mean percentages ± SD of two independent experiments. The activity of each sample was expressed relative to that detected in the empty vector, normalized to 100%. Pair-wise comparison between samples was performed by the Welch’s t-test (unequal variances, two-tailed distribution).

For Western Blot analysis, 5 µg of microsomes were mixed with Laemmli buffer and incubated at room temperature (RT) for 30 minutes. Samples (5 µg per well) were separated on a precast 4-12% NuPAGE™ polyacrylamide gradient gel (Invitrogen). The anti-SPCA2 (Abcam) and anti-GAPDH (Abcam) antibodies were diluted 1:4000 and 1:5000 in 0.1% TBS-Tween™20 (Fisher BioReagents™), respectively.

## RESULTS

### Identification of the missense variant Chr16:84405221

WES analysis in six families selected for SLI led to the identification of a rare variant (Chr16:84405221) in the *ATP2C2* gene (Figure 1). This variant was found in two children and their father, all diagnosed with SLI. The same variant was previously reported in an exome sequencing study performed on 43 unrelated probands affected by severe language impairment (49). *ATP2C2* was previously associated with SLI (29). Sanger sequencing in the other family members showed imperfect segregation as the variant was not transmitted to the other two affected sons (Figure 1). The variant was inherited from the father who also scored below expected (i.e., < 15th %ile) on a standardized expressive language measure. The variant leads to a missense substitution from a valine to a methionine in exon three spanning the ATPase domain. Bioinformatics tools for pathogenicity suggest the change is likely to be benign (Table 1). The two algorithms that predicted a functional effect, albeit subtle, on protein activity and stability were the Mutation Assessor and the INPS-MD tools (Table 1).

### Genotyping of Chr16:84405221 in the SLI, dyslexia and ADHD cohorts

The variant was genotyped in independent cohorts with language impairment, dyslexia, or ADHD (Table 2). The frequency of the variant was 2.2% in the SLIC cohort selected for SLI and 1.8% across all the unrelated LI cases (combined LI samples, N = 360), including the discovery family and previously published data (49). The frequency was 0.8% in the combined dyslexia samples (N = 520), and 0.7% in the combined ADHD cases (N = 151).

In the ALSPAC super-controls (N = 681) the variant occurred at a frequency of 1%. The frequency in the global gnomAD database is of 0.5% (N = 59718) and, of 0.9% in the European population (N = 24046).

Although the frequency was higher in the LI cases (1.5%) than the controls (1%) derived from the ALSPAC cohort, the difference was not statistically significant when examined with a two-tailed Fisher’s exact test (p = 0.152).

### Mean score of language and reading-related phenotypes

We evaluated the effects of the variant on a range of language-related quantitative measures available for the ALSPAC cohort (Supplementary Table S4). The carriers (N = 42) had lower mean scores compared to the non-carriers (N = 1825) on all tested measures, reaching nominal statistical significance (t-test, two-tailed distribution) for six traits: READ (p = 0.016), SPELL (p = 0.006), PHONEME (p = 0.026), NW_READ (p = 0.002), VIQ (p = 0.006) and PIQ (p = 0.022) (Table 3). NW_READ survived a conservative Bonferroni correction for multiple comparison (10 tests, threshold for significance = 0.005; see also Supplementary Table S5 for analysis through the Wald test and Supplementary Figure S2 for box plots). It is worth noting that the 10 tests are not completely independent given the correlation across these measures (83>.

**Table 3.**
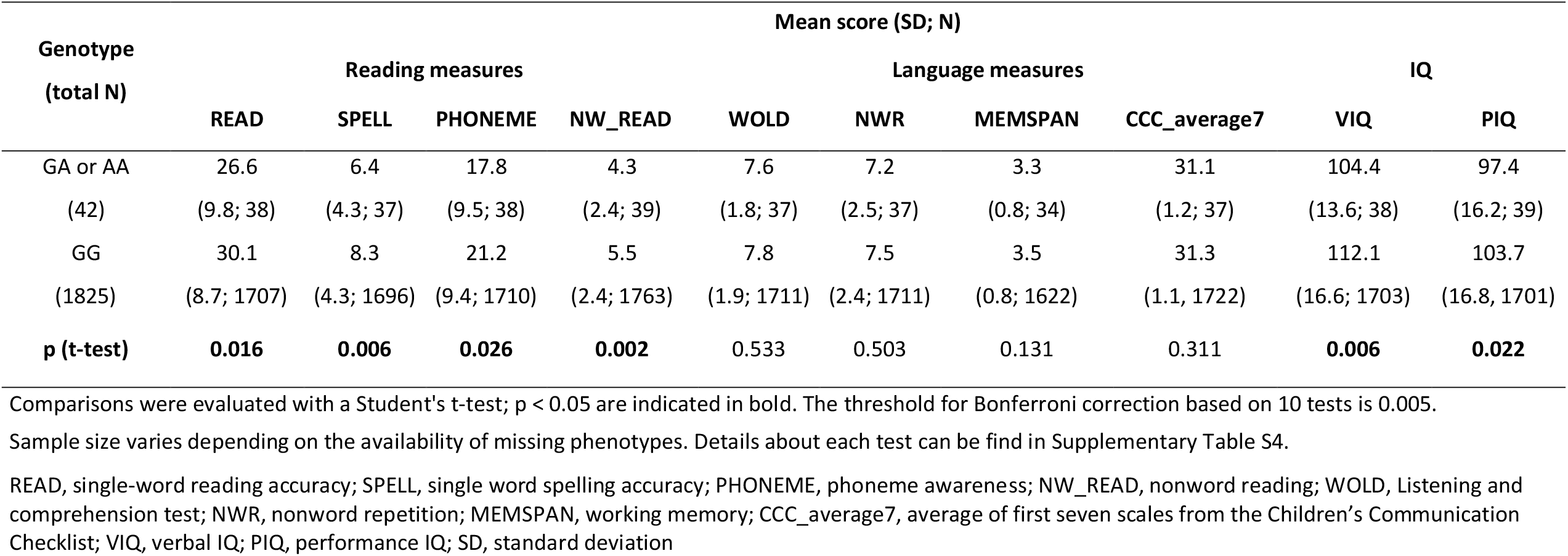
Mean score of different phenotypes across carriers and non-carriers (ALSPAC cohort)

### ATPase assay

An effect of allele A on the ATPase activity of SPCA2 was tested with a colorimetric assay to quantify the inorganic phosphate released during the catalytic reaction (Figure 2A). Microsomes were isolated from HEK293 cells overexpressing the wildtype or mutated SPCA2-HA tagged protein. The activity of the overexpressed SPCA2 WT sample was higher compared to the negative controls (empty vector and untransfected cells) across all the conditions. SPCA2 p.V102M displayed on average a lower ATPase activity compared to that of the WT protein (Figure 2A). WB confirmed the overexpression of SPCA2 for the WT and mutated constructs while no protein was detected for the negative controls, suggesting a very minimal effect of baseline endogenous protein (Figure 2B).

**Figure 2.**
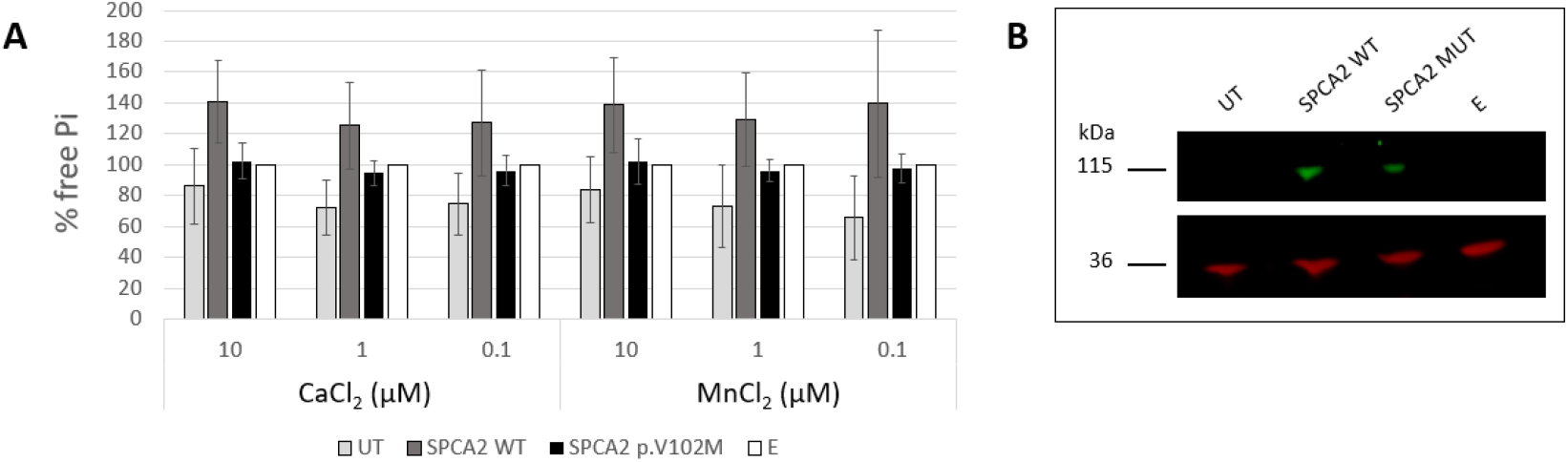
Effect of the mutation on ATPase activity. **A**) Concentration of free inorganic phosphate (Pi) released during the ATPase catalytic reaction. The activity is expressed relative to the empty vector, normalized to 100%. The results are shown as mean percentages ± SD of two independent experiments. The samples are: untransfected HEK293 cells (UT), microsomes isolated from HEK293 cells transiently transfected to overexpress the recombinant SPCA2-HA tagged (SPCA2 WT) or mutated protein (SPCA2 p.V102M) and microsomes isolated from HEK293 cells transiently transfected with the empty vector (E). **B**) Western blot. Each lane was loaded with 5 µg of microsomes. The membranes were probed with antibodies against SPCA2 (top, green) or GAPDH (36 kDa) (bottom, red). SPCA2 can be detected when overexpressed, while no signal is observed in the untransfected cells and the control samples. The band corresponds to the predicted SPCA2-HA tagged molecular weight (105.2 kDa).

## DISCUSSION

In this study we investigated the effects of a rare *ATP2C2* variant on language impairment and related disorders. Overall, we found that this variant had a higher frequency in LI samples (1.8%, N = 360) compared to controls (1%, N = 681, ALSPAC) and subjects with dyslexia (0.8%, N = 520) or ADHD (0.7%, N = 150). The frequency reached 2.2% in a clinical cohort ascertained for SLI (N = 161; Table 2). In a general population sample derived from the ALSPAC cohort, carriers of the rare variants presented lower scores compared to non-carries on a range of language-related measures, with the stronger effect observed for the nonword reading measure (p = 0.002; Table 3). Taken together, our results suggest that the *ATP2C2* missense variant is associated to LI and lower scores on reading and language measures. The presence of the *ATP2C2* variant in controls shows that it is not sufficient on its own to cause a disorder, and its association with a range of measures suggests a pleiotropic effect on cognitive traits. These observations are in line with the emerging findings in the field of complex traits genetics and, specifically, for neurodevelopmental and psychiatric disorders. For example, under the double-hit model, a risk factor might lead to a specific disorder or a range of conditions depending on the interactions with other genetic risk variants (51,52,84>. Under this model, the rare *ATP2C2* variant might increase the risk for LI or affect performance on reading- and language-related measures when combined with different susceptibility factors, as observed for other rare variants implicated in LI. These include a microdeletion in *ZNF277* (42), a rare stop-gain variant in *ERC1* (49) and nonsynonymous mutations in *NFXL1* (85), *GRIN2B* and *SRPX2* (Chen et al., 2017). Increasing evidence shows that the penetrance of rare risk factors is modulated by the cumulative effect of common risk alleles, even in severe neurodevelopmental disorders usually assumed to have a monogenic origin (86). Different manifestations determined by the interplay between common and rare variants have been reported for conditions such as schizophrenia (87,88), ADHD (89), autism spectrum disorder (90), intellectual disability (91) and outcomes such as educational attainment (92). Rare risk factors causing psychiatric disorders have also been reported to have an effect on cognitive abilities in the general population (93,94). This is in line with what we observe for the *ATP2C2* variant which showed associations with poor performance on cognitive traits in the ALSPAC sample regardless of any disorder group assignement (Table 3). The variant is associated to lower mean scores on a range of cognitive measures, with the strongest effect observed for reading abilities rather than language skills. However, although tempting, we should not speculate on specific genotype-phenotype relationship. Instead our results support the emerging idea that genetic risk factors have generalised effects across cognitive and neurodevelopmental traits. The same concept is illustrated also for common variants through polygenic risk scores (PRS) analysis. For example, PRS for schizophrenia showed associations for a number of traits including poor cognitive outcomes (95) but also enhanced creativity (96), language performance (97) and hearing (98). Similarly, polygenic risk scores (PRS) for general cognitive abilities show effects on reading and language measures (15,99–101>.

The *ATP2C2* gene encodes for an ATPase that transports Ca^2+^ and Mn^2+^ into the Golgi lumen (102>. ATPases are emerging as a new class of proteins involved in neurodevelopmental disorders (103– 106). Ca^2+^ is involved in the regulation of several neuronal processes such as neuronal migration, axon guidance, synaptic plasticity, memory, neuronal excitability, neurosecretion and aging (107– 110). Excessive accumulation of Mn^2+^ inside the cells is toxic, induces apoptosis and causes manganism, a neurological disorder characterised by tremors and spasm (111,112). Additionally, dysregulation of these cations has been linked to Alzheimer’s, Parkinson’s and Huntington’s disease (113,114). The transport of Ca^2+^ and Mn^2+^ ions inside the Golgi is essential for the activity of enzymes involved in mechanisms such as neurotransmitter synthesis, membrane trafficking, metabolism, protein modification, sorting and folding (110). Previous studies have shown that *ATP2C2* is expressed in postnatal rat hippocampal neurons (115) and in adult human brain (102,115,116), suggesting a possible role of *ATP2C2* in maintaining ionic homeostasis in neurons.

The missense variant described in the current study is localised in the cation transporting N-terminal domain and causes the amino acid substitution from valine to methionine (Figure 1). The latter residue is characterised by an unbranched side chain that provides increased flexibility. In addition to this, methionine has a sulphur group that can influence the binding preference for metal ions, and be reversibly oxidised converting the hydrophobic amino acid into a highly polar one (117,118>. Bioinformatic predictions do not attribute substantial detrimental effects to the substitution, and therefore any potential effects are expected to be subtle (Table 1). We tested experimentally whether the rare allele A had an effect on the SPCA2 activity, through an ATPase assay. This approach allowed us to measure the hydrolysis of ATP in a quick and sensitive way. Independently of the concentrations tested, the rare variant displayed on average a lower ATPase activity compared to SPCA2 WT (Figure 2). Although not statistically significant, the data suggested a possible impact of the variant on the velocity of the reaction and therefore the cation binding affinity. More specifically, the mutated transporter tended to present a lower ATP hydrolysis indicative of a lower turnover rate and consequently a higher ion affinity. In line with the bioinformatics prediction, the rare variant does not affect the functionality of the protein but might alter its performance which in turn, in combination with other factors, might contribute to neurodevelopmental phenotypes.

In summary, we identified a rare missense variant (chr16:84405221, GRCh38.p12) in *ATP2C2*, a previously reported candidate gene for LI. We showed that the rare allele A increases the risk for LI cohorts and is associated with more general cognitive abilities in a population-based cohort. Preliminary functional characterization of the rare allele suggested that the rare variant influences the ATPase activity of SPCA2 supporting follow up functional studies to fully understand the role of this variant and the ATP2C2 gene. WES studies for language disorders have been limited to samples, which makes the result interpretation particularly challenging. Our results demonstrate that combined analyses across disorders and phenotypes provide a valid route to validate WES findings. Taken together, our results further support a role of *ATP2C2* in language impairment and pinpoint to a specific rare variant which might impact ATPase activity of the SPCA2 protein.

## Data Availability

Data used in this study are available from UK10K Data Access Committee and the ALSPAC Executive Committee for researchers who meet the criteria for access to confidential data.

## Acknowledgement

SP if funded by the Royal Society. This work was supported by an Action Medical Research Action/The Chief Scientist (CSO) Office, Scotland grant (GN2614) and a Cunningham Trust grant to SP and SJP. Support to the analysis was provided by the St Andrews Bioinformatics Unit funded by the Wellcome Trust [grant 105621/Z/14/Z]. Assessment of the Aston cohort was supported by funding from The Waterloo Foundation to JBT and SP [797–1720]. Analysis of the discovery pedigree was supported by the University of Kansas grant (NIH:NIDCD 5 R01 DC001803). Analysis of the York cohort was funded by Wellcome Trust Programme Grant 082036/B/07/Z. DFN is currently supported by Oxford Brookes University funds, the Leverhulme Trust and the British Academy. This work was, in part, completed while DFN was at the Wellcome Trust Centre for Human Genetics, Oxford as an MRC Career Development Fellow (G1000569/1). We would like to thank the members of all teams who collected the data and the families who participated.

We are extremely grateful to all the families who took part in the ALSPAC study, the midwives for their help in recruiting them, and the whole ALSPAC team, which includes interviewers, computer and laboratory technicians, clerical workers, research scientists, volunteers, managers, receptionists and nurses. The UK Medical Research Council and Wellcome (Grant ref: 217065/Z/19/Z) and the University of Bristol provide core support for ALSPAC. This publication is the work of the authors and DFN will serve as guarantors for the analysis of the ALSPAC data presented in this paper. A comprehensive list of grants funding is available on the ALSPAC website (http://www.bristol.ac.uk/alspac/external/documents/grant-acknowledgements.pdf). Whole-genome sequencing of the ALSPAC samples was performed as part of the UK10K onsortium (a full list of investigators who contributed to the generation of the data is available from www.UK10K.org.uk). ALSPAC GWAS data was generated by Sample Logistics and Genotyping Facilities at Wellcome Sanger Institute and LabCorp (Laboratory Corporation of America) using support from 23andMe. We thank Michael Gill for sharing data for the ADHD cohort and Arathi Jeyaratnam for the technical support.

