## Supplementary Information for "A rare missense variant in the *ATP2C2* gene is associated with language impairment and related measures"

### Supplementary materials and methods

#### The ALSPAC cohort

Pregnant women resident in Avon, UK with expected dates of delivery 1st April 1991 to 31st December 1992 were invited to take part in the study. The initial number of pregnancies enrolled is 14,541 (for these at least one questionnaire has been returned or a "Children in Focus" clinic had been attended by 19/07/99). Of these initial pregnancies, there was a total of 14,676 foetuses, resulting in 14,062 live births and 13,988 children who were alive at 1 year of age.

When the oldest children were approximately 7 years of age, an attempt was made to bolster the initial sample with eligible cases who had failed to join the study originally. As a result, when considering variables collected from the age of seven onwards (and potentially abstracted from obstetric notes) there are data available for more than the 14,541 pregnancies mentioned above. The number of new pregnancies not in the initial sample (known as Phase I enrolment) that are currently represented on the built files and reflecting enrolment status at the age of 24 is 913 (456, 262 and 195 recruited during Phases II, III and IV respectively), resulting in an additional 913 children being enrolled. The phases of enrolment are described in more detail in the cohort profile paper and its update (see footnote 4 below). The total sample size for analyses using any data collected after the age of seven is therefore 15,454 pregnancies, resulting in 15,589 foetuses. Of these 14,901 were alive at 1 year of age.

#### Allele-specific PCR

Samples carrying the missense variant when screened via a TaqMan™ SNP Genotyping Assay, were validated by allele-specific PCR. The method implied two forward primers that differ by one base at the

3' end, and a common reverse primer (Supplementary Table S1). Primers were designed using the ApE software (A plasmid Editor, https://jorgensen.biology.utah.edu/wayned/ape/) and the reference *ATP2C2* sequence from the University of California, Santa Cruz (UCSC) genome browser (Human GRCh38/hg38). The optimised PCR conditions and the programme are listed in Supplementary Table S2 and Supplementary Table S3, respectively. The allele-specific forward primers and the common reverse primer were combined in two parallel PCR reactions. Depending on the allele present, the forward primer anneals perfectly to the template or forms a mismatch. The reaction gives an amplicon only in the first case (Supplementary Figure S1).

**Microsome preparation**

At 72 h post-transfection, HEK293 cells were washed twice in ice-cold TBS (Fisher Scientific™) and lysed with 2 ml hypotonic buffer (10 mM Tris-HCl (pH 7.5), 0.5 mM MgCl2) and protease inhibitors (Roche cOmplete™, Mini Protease Inhibitor Cocktail). The cell lysate was then incubated on ice for 10 minutes. An equal volume of 10 mM Tris-HCl (pH 7.3), 0.5 M sucrose and protease inhibitors was added to the samples, and the cells were homogenized with a syringe. The mixture was centrifuged for 10 minutes at 10,000 x g at 4°C. The supernatant was collected and centrifuged for 2 hours at 100,000 x g at 4˚C. The microsomal pellet was resuspended in 0.25 M ice-cold sucrose. Aliquots of the microsomes were snap frozen with liquid nitrogen and stored at -80°C.

Protein concentration was assessed with the Qubit® 2.0 Fluorometer following the manufacturer's protocol (Thermo Fisher Scientific™).

**Western Blot**

Microsomes from HEK293 cells were mixed with Laemmli buffer (pH 6.8) containing 4% SDS, 20% glycerol, 10% β-mercaptoethanol, 0.004% bromphenol blue and 0.125 M Tris-HCl and incubated at room temperature (RT) for 30 minutes. Samples (5 µg per well) were loaded on a precast 4-12% NuPAGE™ polyacrylamide gradient gel (Invitrogen) and run for 2 hours at 190 V using MOPS as running buffer (Invitrogen). Proteins were transferred from the gel to a nitrocellulose membrane (GE Healthcare Amersham™ Protran™, Fisher Scientific) at 360 mA for 2 hours at 4°C. The membrane was blocked for 1 hour at RT with WesternBreeze™ solution (Thermo Fisher Scientific). After blocking, the membrane was incubated overnight at 4°C with the primary antibody. The anti-SPCA2 (Abcam) and anti-GAPDH (Abcam) antibodies were diluted 1/4000 and 1/5000 in 0.1% TBS-Tween™20 (TBST) (Fisher BioReagents™), respectively. This step was followed by 3 washes in 0.1% TBST, and incubation with the secondary fluorescent antibody for 1 hour at room temperature. The IRDye® 800CW Donkey Anti-Rabbit IgG H&L (LI-

COR) and IRDye® 680RD Donkey Anti-Mouse IgG H&L (LI-COR) antibodies were diluted 1/15000 and 1/20000 in 0.01% TBST (Fisher BioReagents™), respectively. After 3 washes in 0.1% TBST, the membrane was allowed to dry and imaged with the Odyssey® CLx instrument (LI-COR, Biosciences). Western Blot visualisation was performed with the Image Studio™ Lite software.

### Supplementary Tables

**Supplementary Table S1. List of primers.**

| Forward primer | Reverse primer | Application | Aim |
|---|---|---|---|
| 5' GTTGCTGACAACAGCGAACCTG 3'<br>5' GTTGCTGACAACAGCGAACCTA 3' | 5' CACTTCTCCTTCCTCATGTGC 3' | PCR | validation of the missense variant in the UK cohorts |
| 5' TGCCTTTCAGAGTGGGAGTC 3' | 5' GGATGCCTTCCATCAACATT 3' | Sanger sequencing | validation of the missense variant in the discovery pedigree |
| 5' GAGAGAAAGCTTGCGCCCGCTCACCATGGTC 3' | 5' GAGAGAGCGGCCGCCACATCTTCAGGGTGCATCTGGACTCTC 3' | RT-PCR | cloning |
| 5' TGACAACAGCGAACCTATGTGGAAGAAATACCTG 3' | 5' CAGGTATTTCTTCCACATAGGTTCGCTGTTGTCA 3' | PCR | site-directed mutagenesis |

**Supplementary Table S2. PCR conditions.**

| Component | Volume (uL) | Final concentration |
|---|---|---|
| Betaine (5 M) | 4 | 1 M |
| Immunobuffer (10 X) | 2 | 1 X |
| $MgCl_2$ (50 µM) | 0.5 | 1.25 µM |
| dNTPs (10 mM) | 0.4 | 0.2 mM |
| Forward primer (10 µM) | 1 | 0.5 µM |
| Reverse primer (10 µM) | 1 | 0.5 µM |
| Immolase™ DNA polymerase | 0.2 | 1.0 units / 50 µL PCR |
| gDNA (50 ng/µL) | 1.5 | 3.75 ng/µL |
| Milli-Q® water | 12 | |
| Total | 20 | |

**Supplementary Table S3. PCR Programme.**

| Step | Temperature (°C) | Time | Number of cycles |
|---|---|---|---|
| Activation | 95 | 10 min | 1 |
| Denaturation | 95 | 30 s | |
| Annealing | 65.4 | 30 s | 30 |
| Extension | 72 | 30 s | |
| Final elongation | 72 | 5 min | 1 |

**Supplementary Table S4. Description of the phenotypic measures (adapted from Scerri et al., 2011).**

| Measure | Summary description | Test | Target age (years) | Reference |
|---|---|---|---|---|
| READ | Single-word reading accuracy | The child was asked to read aloud a series of 48 unconnected words which increased in difficulty. | 7.5 | Rust et al., 1993 |
| SPELL | Single-word spelling accuracy | The child was asked to spell a series of 15 regular and irregular words of increasing difficulties. | 7.5 | Nunes et al. 2003 |
| PHONEME | Phoneme awareness | The phoneme deletion task (Auditory Analysis Test) comprised 2 practice and 40 test items of increasing difficulty. The task involved asking the child to repeat a word and then to say it again but with part of the word (a phoneme or number of phonemes) removed. | 7.5 | Rosner et al., 1971 |
| NW_READ | Single-non-word reading accuracy | This was assessed by asking the child to read out loud ten real words, followed by ten non-words. | 9.5 | Nunes et al. 2003 |
| WOLD | Listening and comprehension test | The child was read a paragraph about a picture, which the child is shown. The child then answers questions on what he/she has heard. The child has to make inferences about what was read to him/her and answer the questions verbally. The task was discontinued if the child got three consecutive questions incorrect. | 8.5 | Rust, 1996 |
| NWR | Phonological short-term memory test | An adaptation of the Nonword Repetition Test was used. This comprised twelve nonsense words, four each of 3, 4 and 5 syllables and conforming to English rules for sound combinations. The child was asked to listen to each word via an audio cassette recorder and then repeat each item. | 8.5 | Gathercole et al., 1994 |

| | | | | |
|---|---|---|---|---|
| MEMSPAN | Working memory | Working memory was tested using the Counting Span Task, which requires the simultaneous processing and storage of information. On the computer monitor the child was presented with a number of red and blue dots on a white screen. The child was asked to point to and count the number of red dots out loud (the processing component). After each set, the child was asked to recall the number of red dots seen on each screen in the order they were presented within that set (the storage component). | 10.5 | Case et al., 1982 |
| CCC_average7 | Average of first seven scales from Children's Communication Checklist | The CCC consists of 70 items grouped into 9 subscales with scores defined for each subscale as well as a summary score for pragmatic aspects of communication as the sum of the 3rd to 7th subscales. In this questionnaire the first 53 items making up the first 7 subscales were used. | 7.5 | Bishop, 1998 |
| VIQ<br><br>PIQ | Verbal IQ<br><br>Performance IQ | VIQ and PIQ were assessed using the Wechsler Intelligence Scale for Children - Third Edition (WISC-III). The test comprises five verbal subtests (Information, Similarities, Arithmetic, Vocabulary, Comprehension) and five performance subtests (picture completion, coding, picture arrangement, block design, object assembly). | 8 | Wechsler et al., 1992 |

**Supplementary Table S5. Association of the rare allele A with language and reading-related traits (Wald test statistics) (ALSPAC cohort, N = 1867 individuals with available Chr16:84405221 genotype).**

| Phenotype | N | p-value | β | S.E. |
|---|---|---|---|---|
| READ | 1745 | **0,016** | -3,466 | 1,436 |
| SPELL | 1733 | **0,006** | -1,942 | 0,710 |
| PHONEME | 1748 | **0,025** | -3,442 | 1,539 |
| WOLD | 1748 | 0,533 | -0,198 | 0,318 |
| NWR | 1748 | 0,503 | -0,272 | 0,407 |
| NW_READ | 1802 | **0,002** | -1,207 | 0,396 |
| MEMSPAN | 1656 | 0,131 | -0,217 | 0,144 |
| CCC_average7 | 1759 | 0,280 | -0,204 | 0,189 |
| VIQ | 1741 | **0,005** | -7,692 | 2,710 |
| PIQ | 1740 | **0,022** | -6,222 | 2,713 |

Abbreviations: READ, single-word reading accuracy; SPELL, single word spelling accuracy; PHONEME, phoneme awareness; WOLD, Wechsler Objective Language Dimensions; NWR, nonword repetition; NW_READ, nonword reading; MEMSPAN, memory span; CCC, Children's Communication Checklist; VIQ, verbal IQ; PIQ, performance IQ; N, number of individuals with available information; β= effect size, negative trend indicates that the rare allele is a risk factor; S.E. = Standard Error.

### Supplementary Figures

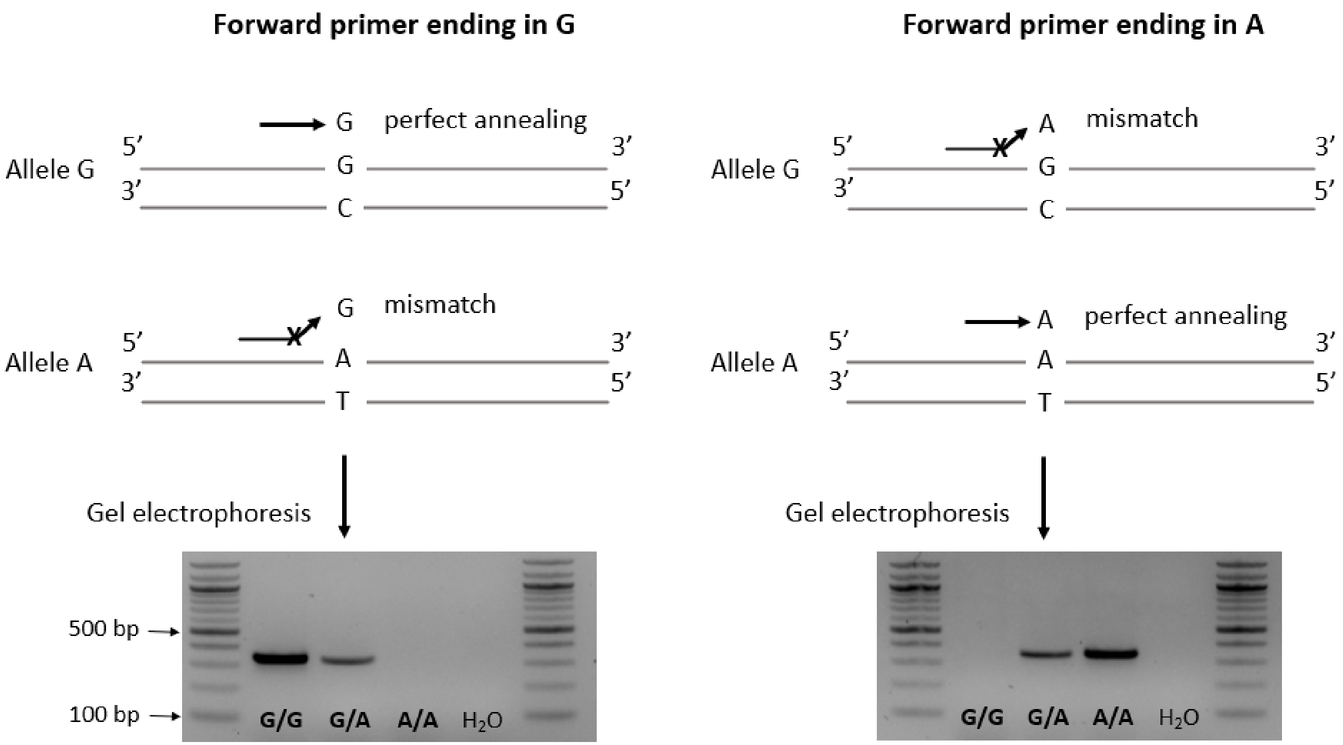


**Supplementary Figure S1: Allele-specific PCR.** Schematic of the DNA amplification in the presence of the forward primer ending in G (on the left) or A (on the right). The reaction is allowed only if the nucleotide at the 3'-end of the primer is complementary to the base of the DNA template**.** The genotype of each sample was determined loading the PCR products on a 2% agarose gel.

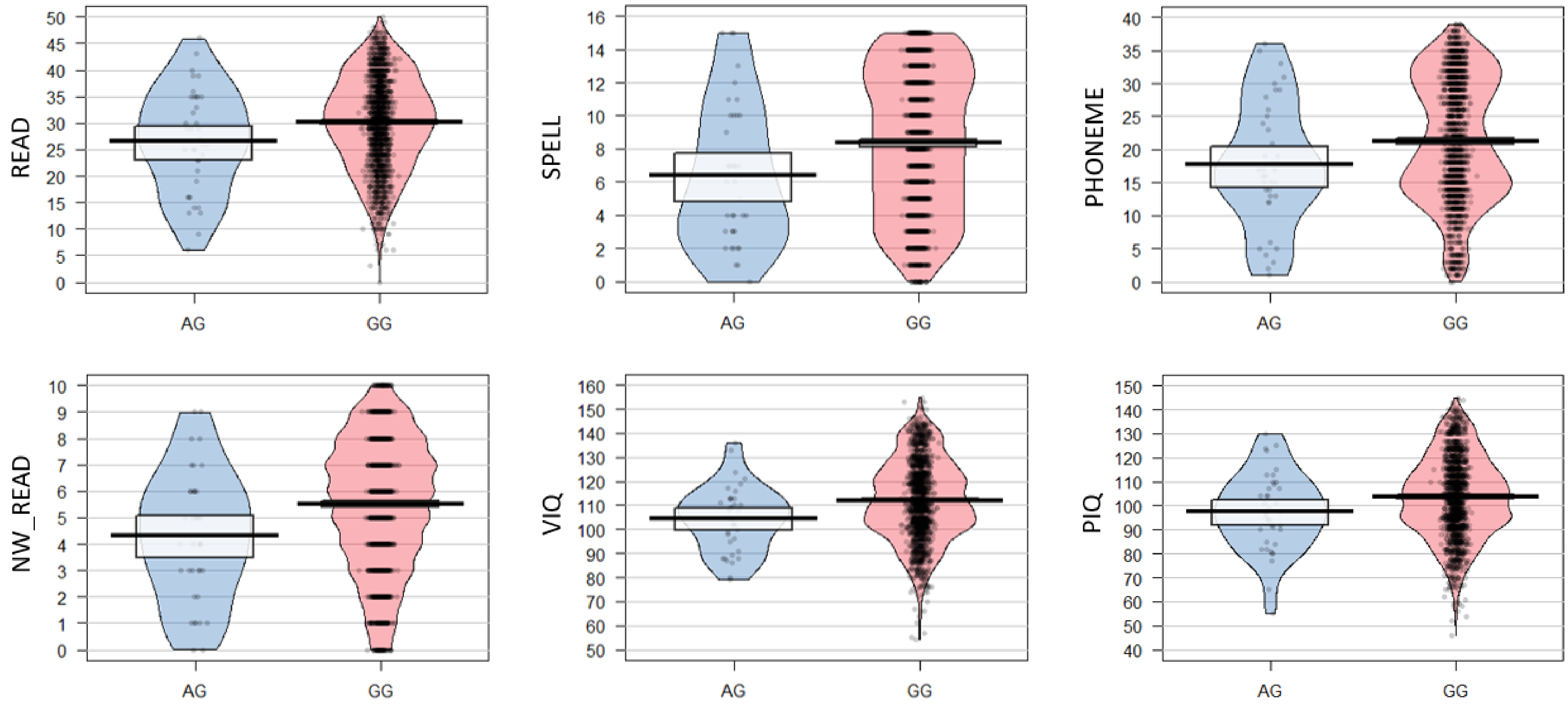


**Supplementary Figure S2: Box plots of the phenotypes found to be associated with the rare missense *ATP2C2* variant (Chr16:84405221).** The charts show the distribution of the different language and reading-related traits in carriers (AG) and non-carriers (GG) (ALSPAC cohort, N = 1867 individuals with available genotype).
